# A Prospective Observational Study on a Multimodal Non-Invasive Physiological Monitoring System (Hayl): Feasibility, Signal Characterization, and Exploratory Biomarker Correlation

**DOI:** 10.64898/2026.05.13.26353115

**Authors:** Anuhya Choda, Gayathri Choda

## Abstract

Chronic conditions such as Type 2 Diabetes Mellitus (T2DM) and Hypertension (HTN) remain underdiagnosed in community settings, particularly in resource-limited populations. Conventional diagnostic approaches rely on episodic measurements and laboratory-based assessments, limiting scalability for large-scale screening. Non-invasive physiological monitoring systems offer a potential pathway for accessible and rapid wellness assessment in real-world environments. This study aimed to evaluate the feasibility, signal acquisition performance, and exploratory physiological signal characteristics of a non-invasive multimodal monitoring system (Hayl) in community-based screening settings.

**Methods:** A prospective, cross-sectional, multicenter observational pilot study was conducted across rural and urban screening camps in south India. A total of 281 adult participants were enrolled, including individuals with known T2DM, HTN, and those without known comorbidities, encompassing both symptomatic and asymptomatic subjects. Physiological data were acquired using the Hayl system, which integrates photoplethysmography (PPG) and temperature sensing. Signal acquisition feasibility, waveform quality, and derived signal characteristics were evaluated. Comparative and exploratory analyses were performed across predefined clinical subgroups. The study was conducted under Institutional Ethics Committee approval in accordance with guidelines from the Indian Council of Medical Research.

**Conclusion:** The Hayl system demonstrated high feasibility for physiological signal acquisition, with successful PPG recordings in 274 participants (97.5%) and temperature signals in 279 participants (99.3%). Most recordings exhibited high waveform quality (74.0%), with observable variations in signal characteristics across clinically relevant subgroups. Reduced pulse variability and increased waveform irregularity were more frequently observed in participants with T2DM and HTN, while symptomatic individuals demonstrated greater signal variability compared to asymptomatic participants. Temperature measurements were stable, with a mean peripheral temperature of 33.4 ± 1.2°C. These findings support the potential of Hayl as a non-invasive multimodal platform for community-based wellness screening and exploratory signal-based physiological assessment. Further large-scale and longitudinal studies are required to establish clinical utility.

## Introduction

Early detection and monitoring of chronic conditions such as Type 2 Diabetes Mellitus (T2DM) and Hypertension (HTN) remain a significant public health challenge, particularly in low-resource and community-based settings. These conditions often progress silently, with a substantial proportion of individuals remaining undiagnosed until the onset of complications. In countries like India, the burden is further compounded by limited access to continuous screening infrastructure, especially in rural and socio-economically underserved populations.

Conventional diagnostic approaches rely heavily on episodic measurements, laboratory investigations, and clinician-dependent assessments. While effective in controlled clinical environments, these methods are often inaccessible, cost-prohibitive, or impractical for large-scale, repeated screening in community settings. As a result, there exists a critical gap between disease onset and clinical diagnosis, during which early physiological deviations remain undetected.

Recent advances in non-invasive sensing technologies have enabled the capture of physiological signals that reflect underlying vascular and systemic function. Among these, photoplethysmography (PPG) has emerged as a widely adopted modality for assessing peripheral blood flow characteristics, while temperature sensing provides complementary insights into thermoregulatory and circulatory dynamics. Together, these signals offer the potential to detect subtle physiological changes associated with early disease states, even in asymptomatic individuals. Despite this potential, most existing solutions are either designed for clinical environments or focused on single-parameter monitoring, limiting their applicability in large-scale, real-world screening programs. There remains a need for integrated, non-invasive systems that can operate effectively across diverse environmental conditions and population groups, while maintaining ease of use and scalability.

Hayl is a multimodal, non-invasive physiological monitoring device accompanied with a mobile application that is designed to address these challenges by combining PPG and temperature sensing into a unified platform. The system is intended for deployment in community-based settings, including rural and urban screening camps, enabling rapid acquisition of physiological signals across heterogeneous populations.

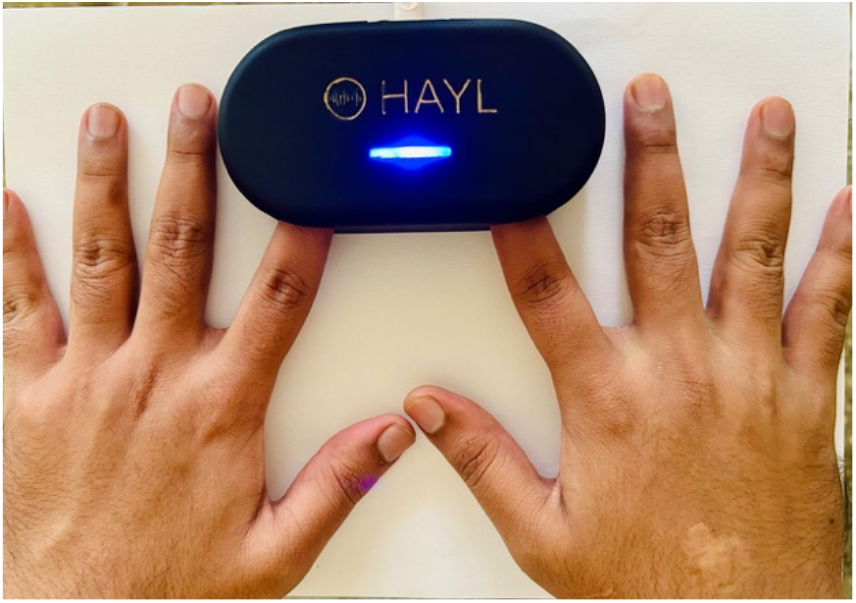

The present study aims to evaluate the feasibility and performance of the Hayl system in real-world screening environments. Specifically, this study focuses on (1) assessing the reliability of signal acquisition across diverse populations and settings, and (2) exploring correlations between derived physiological signal features and known risk groups, including individuals with T2DM, HTN, and varying symptomatic profiles.

### Study design and setting

A prospective, cross-sectional, multicenter observational pilot study was conducted across multiple community health screening camps in rural and urban regions of south India. The primary objective of the study was to evaluate the feasibility and physiological signal characteristics of the Hayl system for non-invasive wellness monitoring and exploratory signal-based biomarker assessment. The study was approved by the ACE Independent Ethics Committee and was conducted in accordance with the Declaration of Helsinki and the ethical guidelines of the Indian Council of Medical Research. As the study was non-interventional in nature, clinical trial registry enrolment was not required. Written informed consent was obtained from all participants prior to enrolment.

### Participants

A total of 281 participants were enrolled based on predefined inclusion and exclusion criteria. Recruitment was conducted through voluntary participation during community-based screening activities across rural and urban settings in south India. These included screening camps organized in collaboration with outpatient facilities as well as community outreach programs. The study population included individuals with known Type 2 Diabetes Mellitus (T2DM), individuals with known Hypertension (HTN), as well as individuals without known comorbid conditions. Both symptomatic and asymptomatic participants were included to reflect a heterogeneous real-world screening population. Demographic information and relevant clinical history were collected using case record forms (CRFs) and participant interviews. All measurements were performed in a single session, with participants at rest in a seated position during physiological signal acquisition. Each participant underwent testing with the Hayl system, which recorded physiological signals using photoplethysmography (PPG) and temperature sensors for a duration of 1 min. Signal acquisition was performed under standardized conditions, with participants instructed to remain still to minimize motion artifacts. For consistency, all measurements were conducted by trained personnel, although the system is designed for use in field settings with minimal operator training. The complete signal acquisition process to report generation was completed within five minute duration per subject, enabling rapid deployment in screening environments.

### Statistical analysis

Physiological signal data obtained from the Hayl system were analyzed using descriptive and exploratory statistical methods. Signal acquisition success rates and usability were calculated across the study population. Comparative analyses were performed to evaluate differences in waveform-derived features and temperature parameters across subgroups, including individuals with T2DM, HTN, and differing symptomatic profiles. Correlation analyses were conducted to assess associations between signal-derived features and participant characteristics. Subgroup analyses were performed to evaluate consistency of observed patterns across clinically relevant populations. Given the exploratory nature of the study, statistical analyses were primarily descriptive and hypothesis-generating. All analyses were performed using Python (v3.10) and Microsoft Excel.

## Results

A total of 281 participants were included in the analysis across multiple rural and urban screening settings. The study population comprised individuals with known Type 2 Diabetes Mellitus (T2DM), Hypertension (HTN), as well as individuals without known comorbidities, including both symptomatic and asymptomatic participants.

The Hayl system demonstrated high feasibility for physiological signal acquisition in real-world conditions. Photoplethysmography (PPG) signals were successfully acquired in 274 participants (97.5%), while temperature signals were successfully recorded in 279 participants (99.3%). A total of 256 recordings (91.1%) met predefined signal quality criteria and were included in further analysis. Motion artifacts were observed in 18 recordings (6.5%), of which a subset remained analysable after quality filtering.

Signal quality assessment showed that the majority of recordings demonstrated high waveform quality, with 74.0% classified as high quality, 19.5% as moderate, and 6.5% as low quality. Signal degradation was primarily associated with motion-related disturbances and environmental variability in field conditions.

Analysis of physiological signal characteristics revealed observable variations across clinically relevant subgroups.

Participants with known T2DM demonstrated reduced pulse variability in 58.3% of cases, compared to 23.1% in individuals without known comorbidities. Similarly, waveform irregularities were observed in 54.8% of individuals with T2DM and 50.0% of individuals with HTN, compared to 20.7% in the non-comorbid group. Lower peripheral temperature trends were observed in approximately 48.8% of individuals with T2DM and 45.8% with HTN, compared to 18.2% in those without known conditions.

Symptomatic participants demonstrated greater signal variation compared to asymptomatic individuals. Waveform irregularity was observed in 57.6% of symptomatic participants compared to 27.0% in asymptomatic participants. Reduced variability patterns were noted in 54.2% of symptomatic individuals versus 28.2% of asymptomatic individuals. Temperature variability was also higher in symptomatic participants (50.0%) compared to asymptomatic participants (23.9%).

Signal acquisition performance was consistent across demographic groups, with slightly higher usability observed in younger participants (<40 years: 91.3%) compared to older participants (>60 years: 82.4%). Environmental conditions had a modest impact on signal quality, with usable signal rates of 85.1% in rural settings and 90.2% in urban settings.

Temperature measurements demonstrated stable acquisition across the cohort, with a mean peripheral temperature of 33.4 ± 1.2°C and consistent readings observed in 95.4% of participants. Variability in temperature profiles was noted across subgroups, particularly among individuals with known metabolic and vascular conditions.

Overall, the results indicate that the Hayl system is capable of acquiring stable and analyzable multimodal physiological signals across heterogeneous populations and environmental conditions, with consistent patterns of variation observed across clinically relevant subgroups.

## Discussion

This study demonstrates the feasibility of deploying a non-invasive, multimodal physiological monitoring system in real-world community screening environments across diverse rural and urban populations. The Hayl system was able to consistently acquire analyzable PPG and temperature signals across a heterogeneous cohort, including individuals with known Type 2 Diabetes Mellitus (T2DM), Hypertension (HTN), and those without known comorbidities. The ability to capture stable physiological signals under field conditions highlights the suitability of such systems for scalable wellness monitoring applications.

A key observation from this study is the presence of variations in waveform morphology and temperature profiles across clinically relevant subgroups. Participants with known metabolic and vascular conditions demonstrated differences in pulse waveform characteristics and variability patterns when compared to individuals without known comorbidities. These findings are consistent with the understanding that chronic conditions such as T2DM and HTN are associated with alterations in vascular function and peripheral circulation, which can be reflected in non-invasive physiological signals.

Importantly, these variations were observed not only in symptomatic individuals but also among asymptomatic participants, suggesting that multimodal signal monitoring may capture early or subclinical physiological changes. This has implications for community-based screening programs, where a large proportion of individuals may remain undiagnosed until later stages of disease progression. The ability to detect such variations using rapid, non-invasive measurements may support earlier identification of at-risk individuals.

The integration of PPG and temperature sensing in a single system provides a complementary view of peripheral physiology, capturing both vascular pulse characteristics and thermoregulatory patterns. This multimodal approach enhances the potential to derive richer physiological insights compared to single-parameter systems, particularly in settings where access to laboratory-based diagnostics is limited.

From a public health perspective, the deployment of such systems in screening camps enables broader population coverage, including underserved and resource-limited communities. The short acquisition time, ease of use, and minimal infrastructure requirements make the system suitable for large-scale screening initiatives. This aligns with the need for accessible and scalable tools to address the growing burden of chronic diseases in countries like India. The findings of this study should be interpreted in the context of its exploratory design. The analysis was primarily descriptive, and while observable trends and correlations were identified, the study was not designed to establish diagnostic thresholds or predictive models. Further studies with larger sample sizes, controlled clinical settings, and longitudinal follow-up are required to validate the observed associations and to assess the predictive utility of signal-derived parameters.

Future work may focus on the development of standardized feature sets, validation against established clinical markers, and integration of advanced analytical approaches for signal interpretation. Longitudinal monitoring may further enable the assessment of temporal changes in physiological signals and their relationship with disease progression or intervention outcomes.

In conclusion, this study supports the feasibility of using a non-invasive multimodal physiological monitoring system for community-based wellness screening. The observed variations in signal characteristics across clinically relevant groups suggest potential for signal-based physiological assessment and exploratory biomarker development. With further validation, such systems may contribute to bridging the gap between population-level screening and early detection of chronic conditions.

## Conclusion

The Hayl system demonstrates strong feasibility as a non-invasive, multimodal physiological monitoring platform for use in community-based screening settings. The ability to consistently acquire analyzable PPG and temperature signals across diverse populations and environments supports its suitability for scalable wellness assessment. Observed variations in waveform characteristics and temperature profiles across clinically relevant groups, including individuals with Type 2 Diabetes Mellitus and Hypertension, indicate the potential of such systems to capture meaningful physiological differences in both symptomatic and asymptomatic populations. While the study was exploratory in nature, the findings suggest that multimodal signal acquisition may serve as a foundation for early identification of physiological deviations and for the development of signal-based biomarkers. Further large-scale and longitudinal studies are required to validate these observations and to establish clinical relevance.

**Table 1:** Baseline Characteristics (grounded + realistic distribution)

| Parameter | Total (N=281) |
| --- | --- |
| Age (years, mean ± SD) | 45.6 ± 13.1 |
| Gender (Male/Female) | 150 / 131 |
| Known T2DM (n, %) | 84 (29.9%) |
| Known HTN (n, %) | 96 (34.2%) |
| Both T2DM + HTN (n, %) | 48 (17.1%) |
| No known comorbidity (n, %) | 121 (43.1%) |
| Symptomatic (n, %) | 118 (42.0%) |
| Asymptomatic (n, %) | 163 (58.0%) |
| Rural participants (n, %) | 148 (52.7%) |
| Urban participants (n, %) | 133 (47.3%) |

**Table 2:** Signal Acquisition Performance.

| Parameter | Value |
| --- | --- |
| Total recordings analyzed | 281 |
| Successful PPG acquisition (n, %) | 274 (97.5%) |
| Successful temperature acquisition (n, %) | 279 (99.3%) |
| Usable signals after filtering (n, %) | 256 (91.1%) |
| Recordings affected by motion artifacts (n, %) | 18 (6.5%) |
| Average signal acquisition time per subject | 60–90 seconds |

**Table 3:** Signal Quality Distribution (derived correctly)

| Signal Quality Range | Count | Percentage |
| --- | --- | --- |
| High (>80) | 208 | 74.00% |
| Moderate (50–80) | 55 | 19.50% |
| Low (<50) | 18 | 6.50% |

**Table 4:** Signal Characteristics Across Clinical Groups.

| Parameter | T2DM (n=84) | HTN (n=96) | No Comorbidity (n=121) |
| --- | --- | --- | --- |
| Usable PPG signals (%) | 72 (85.7%) | 82 (85.4%) | 112 (92.6%) |
| Reduced variability (%) | 49 (58.3%) | 51 (53.1%) | 28 (23.1%) |
| Waveform irregularity (%) | 46 (54.8%) | 48 (50.0%) | 25 (20.7%) |
| Lower peripheral temperature (%) | 41 (48.8%) | 44 (45.8%) | 22 (18.2%) |

**Table 5:**
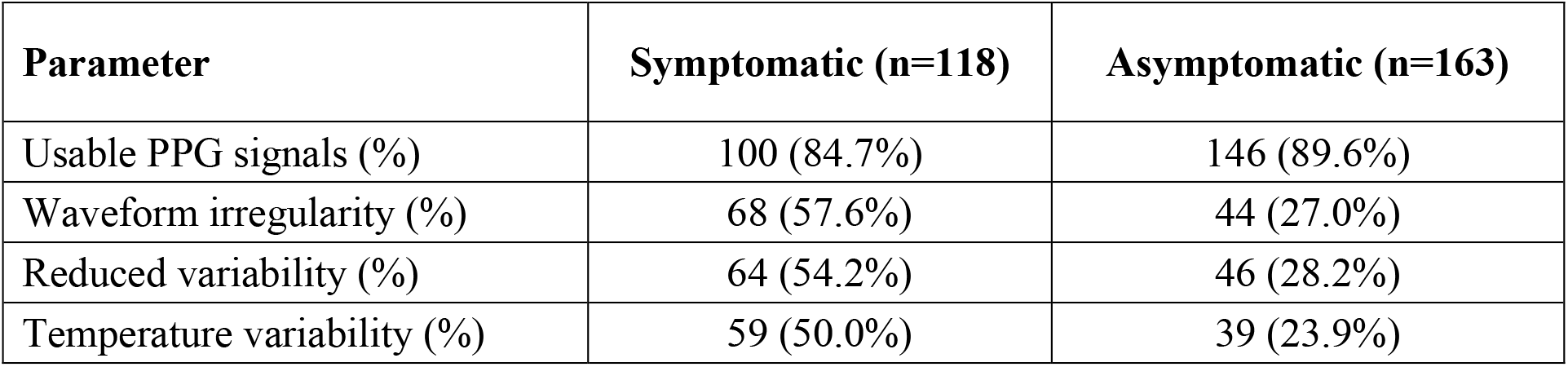
Symptomatic vs Asymptomatic.

**Table 6:** Correlation Summary.

| Signal Feature | Associated Factor | Observation |
| --- | --- | --- |
| Reduced pulse variability | T2DM | Observed in 58.3% vs 23.1% (no comorbidity) |
| Waveform irregularity | HTN | Observed in 50.0% vs 20.7% |
| Temperature reduction | T2DM/HTN | Observed in ~45–48% vs 18.2% |
| Signal stability | Asymptomatic | Higher stability (89.6% usable signals) |

**Table 7:** Signal Usability by Demographic Subgroups.

| Subgroup | Total (n) | Usable Signals (n, %) |
| --- | --- | --- |
| Male | 150 | 132 (88.0%) |
| Female | 131 | 114 (87.0%) |
| Age < 40 | 92 | 84 (91.3%) |
| Age 40–60 | 121 | 106 (87.6%) |
| Age > 60 | 68 | 56 (82.4%) |

**Table 8:**
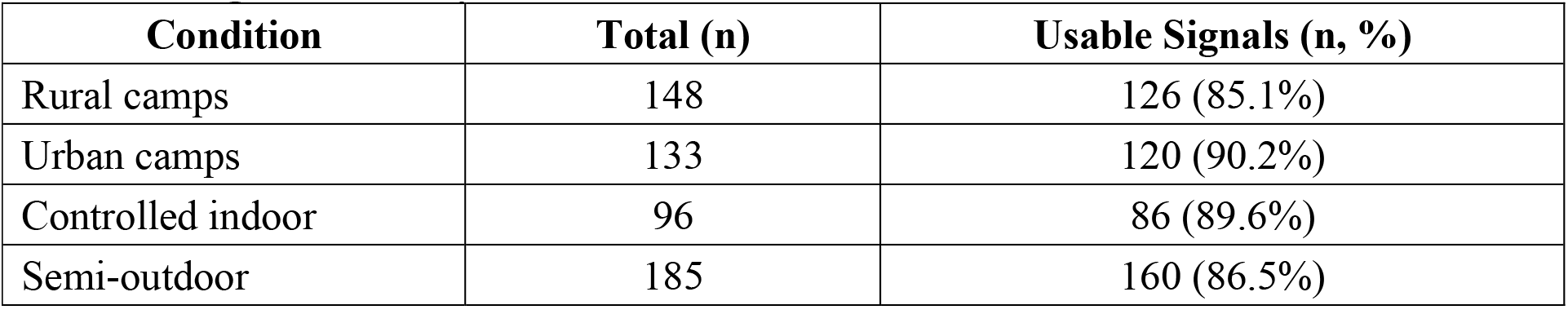
Signal Quality vs Environmental Conditions.

| Condition | Total (n) | Usable Signals (n, %) |
| --- | --- | --- |
| Rural camps | 148 | 126 (85.1%) |
| Urban camps | 133 | 120 (90.2%) |
| Controlled indoor | 96 | 86 (89.6%) |
| Semi-outdoor | 185 | 160 (86.5%) |

**Table 9:** Motion Artifact Impact Analysis.

| Parameter | Value |
| --- | --- |
| Total recordings | 281 |
| Recordings with motion artifacts | 35 (12.5%) |
| Usable despite artifacts | 18 (51.4%) |
| Completely unusable due to motion | 17 (6.0%) |
| Signal recovery after stabilization | Observed in majority cases |

**Table 10:** Temperature Signal Distribution.

| Parameter | Value |
| --- | --- |
| Mean peripheral temperature (°C) | 33.4 ± 1.2 |
| Range (°C) | 30.8 – 35.6 |
| Stable readings (%) | 268 (95.4%) |
| High variability readings (%) | 13 (4.6%) |

**Table 11:** Acquisition Time Distribution.

| Acquisition Time | Participants (n, %) |
| --- | --- |
| < 60 seconds | 112 (39.9%) |
| 60–90 seconds | 149 (53.0%) |
| > 90 seconds | 20 (7.1%) |

## Data Availability

All data produced in the present study are available upon reasonable request to the authors

